# Effect of Proton Pump Inhibitor Deprescription on the Risk of *Clostridioides difficile* Infection: A Quasi-Interventional Substudy of MedSafer

**DOI:** 10.64898/2026.07.22.26358694

**Authors:** Connor Prosty, Yohann Pilon, Jimin J Lee, Todd C. Lee, Emily G. McDonald

## Abstract

**Background:** Proton pump inhibitors (PPIs) have been proposed as a risk factor for initial and recurrent *Clostridioides difficile* infection (CDI) based on observational data. Whether desprescribing PPIs reduces the odds of incident CDI is unknown.

**Methods:** This was a secondary analysis of the cluster randomized MedSafer clinical trial dataset, in which hospitalized older adults taking five or more medications were randomized to usual care (a medication review) or electronic deprescribing decision support prior to hospital discharge. For this substudy, all patients were included regardless of trial assignment and were categorized according to their PPI use at hospitalization and discharge. The 30-day occurrence of CDI was compared as a function of PPI use using a Bayesian mixed-effect logistic regression adjusted for prespecified confounders.

**Results:** 4990 patients were included in the analysis, including 405 (8.1%) patients who were deprescribed their home PPI and 325 (6.5%) new PPI users. 30-day CDI occurred in 32 (0.6%) patients. PPI deprescription was associated with a 99.1% probability of increased odds of CDI (adjusted odds ratio[aOR]=3.66, 95%Credible interval[95%CrI]=1.25-10.11). New PPI use, however, was not associated with a probable increase in CDI (aOR=1.14, 95%CrI=0.20-4.67).

**Conclusion:** Unexpectedly, PPI deprescription was associated with a higher odds of CDI. This challenges the practice of deprescribing unnecessary PPIs in hospitalized patients for CDI prevention. However, it is possible this finding is artefactual due to confounding by indication. Further mechanistic and confirmatory studies are needed.

**SOCIAL MEDIA POST:** In a secondary analysis of a recent deprescribing software trial, proton pump inhibitor deprescription but not new use increased the odds of 30-day *Clostridioides difficile*.

## INTRODUCTION

*Clostridioides difficile* infection (CDI) is a common cause of healthcare- and community-acquired diarrheal infection. The pathogenesis of CDI is thought to be *C. difficile* colonization and a disrupted gut microbiome, most commonly by systemic antibiotics^1^. Proton pump inhibitors (PPIs) are also frequently identified as a risk factor for incident and recurrent CDI in observational studies^2,3^, which is postulated to be from microbiome modulation or increased susceptibility to *C. difficile* colonization^4^. However, there is a paucity of interventional data supporting this link^5^. Current CDI guidelines do not specifically position on PPI deprescription in patients with CDI^6,7^. In a nationwide French study, 30% of adults were dispensed at least one PPI prescription in 2015^8^. Further, 52.9% of patients with CDI on a continuous PPI in one study lacked an evidence-based indication for such use^2^. Given the prevalence and potential harm associated with unnecessary PPI use^9^, deprescription of unnecessary PPIs may offer a simple strategy to mitigate CDI risk. This is particularly true among older adults who are frequently on long-term PPIs without guideline-supported indications^2^ and are at higher risk for CDI.

The MedSafer trial was a cluster randomized controlled trial (RCT) that compared the efficacy of an electronic deprescribing decision support tool versus usual care on adverse drug events after hospital discharge in older adults (≥65 years) with polypharmacy (five or more usual home medications)^10^. PPIs were one of the most commonly identified potentially inappropriate medications for which deprescription was recommended. Using this data, we sought to determine the effect of PPI deprescription on CDI.

## METHODS

### Design and Population

This was a *post hoc* secondary analysis of the MedSafer RCT dataset^10^. The entire study population was included, regardless of assignment, because PPIs were deprescribed in both the intervention and control groups^10^. The analysis plan was pre-specified.

### Exposure

Exposure status was ascertained at hospital discharge and was divided into four categories: 1) patients on a home PPI that was deprescribed (PPI deprescribed), 2) patients on a home PPI that was continued (PPI continued), 3) patients not on a home PPI who were started on one (new PPI users), and 4) patients who did not use a PPI at admission or during hospitalization (PPI non-users).

### Outcome

The primary outcome was CDI within 30 days of discharge. As a counter-balancing measure of harm from deprescribing PPIs, a secondary outcome was upper gastrointestinal bleeding (UGIB) at day 30^10^. Both outcomes were established by patient report during a structured interview on day 30^10^ and, whenever possible, validated by chart review.

### Statistical Analysis

A Bayesian approach was employed because it provides a more nuanced, probabilistic interpretation of evidence. Analyses were performed in R (R Core Team, 2024, Vienna, Austria) using the *brms* package^11^. We fitted a Bayesian mixed-effects multivariable logistic regression model, using a Bernoulli likelihood, with CDI or UGIB as the outcome and PPI exposure as the predictor. Four Markov chains were run with 10,000 iterations, including a 2,000 iteration warm-up. We used the weakly informative Cauchy priors for the predictors (0, 2.5) and intercept (0,10) recommended by Gelman et al.^12^. Cluster was included as a random effect to account for regional variation in CDI. Results were computed as an adjusted odds ratio (aOR) with a 95% credible interval (95%CrI). The probability of any benefit or harm from the PPI exposure category was calculated from the posterior distribution. The model was adjusted for prespecified confounders, including systemic antibiotic use at discharge, prior CDI, and age for the CDI outcome, as well as age, cirrhosis, and anticoagulant/antiplatelet use at discharge for the UGIB outcome.

## RESULTS

### Demographics

The 4990 patient MedSafer cohort was 50.6% female and the median age was 78 years (**Table 1)**. Among the total cohort, 7.1% and 65.3% had systemic antibiotic and anticoagulant/antiplatelet use at discharge, respectively, and 5.6% had cirrhosis. There was history of CDI and UGIB preceding or during the current admission in 2.8% and 6.5%, respectively. According to the PPI exposure categories, 405 (8.1%) were PPI deprescribed, 1957 (39.2%) were PPI continued, 325 (6.5%) were new PPI users, and 2303 (46.2%) were PPI non-users.

**Table 1.** Demographic data.

| Variable | PPI Deprescribed<br>N (%) | PPI Continued<br>N (%) | New PPI Users<br>N (%) | PPI Non-users<br>N (%) | Total<br>N (%) |
| --- | --- | --- | --- | --- | --- |
| Total N | 405 | 1957 | 325 | 2303 | 4990 |
| Median Age (IQR) | 80 (73, 87) | 78 (71, 84) | 77 (71, 83) | 79 (72, 86) | 78 (72, 85) |
| Female Sex | 214 (52.8%) | 1045 (53.4%) | 142 (43.7%) | 1123 (48.8%) | 2524 (50.6%) |
| Cirrhosis | 22 (5.4%) | 121 (6.2%) | 18 (5.5%) | 68 (3.0%) | 229 (4.6%) |
| Anticoagulant or antiplatelet use at discharge | 248 (61.2%) | 1333 (68.1%) | 211 (64.9%) | 1468 (63.7%) | 3260 (65.3%) |
| Systemic antibiotics at discharge | 23 (5.7%) | 153 (7.8%) | 27 (8.3%) | 153 (6.6%) | 356 (7.1%) |
| Prior CDI (including during the present admission) | 13 (3.2%) | 61 (3.1%) | 9 (2.8%) | 58 (2.5%) | 141 (2.8%) |
| Prior UGIB (including during the present admission) | 26 (6.4%) | 180 (9.2%) | 84 (25.8%) | 36 (1.6%) | 326 (6.5%) |
| 30-day CDI post-discharge | 7 (1.7%) | 9 (0.5%) | 2 (0.6%) | 14 (0.6%) | 32 (0.6%) |
| 30-day UGIB post-discharge | 3 (0.7%) | 8 (0.4%) | 7 (2.2%) | 12 (0.5%) | 30 (0.6%) |

### 30-Day CDI

A total of 32 (0.6%) patients experienced CDI within 30 days of discharge, 14 (44%) of which were recurrences (defined as any preceding episode of CDI) . Compared to continued PPI use, deprescription was associated with a 99.1% probability of increased CDI odds (aOR=3.66, 95%CrI=1.25-10.11), whereas no highly probable association was detected for non-users (aOR=1.38, 95%CrI=0.62-3.22) or new users (aOR=1.14, 95%CrI=0.20-4.67, **Table 2**).

**Table 2.** Mixed-effects Bayesian logistic regression for 30-day CDI and UGIB.

| Predictor | CDI<br>aOR (95%CrI) | UGIB<br>aOR (95%CrI) |
| --- | --- | --- |
| PPI Deprescribed | 3.65 (1.25-10.11) | 1.83 (0.43-6.38) |
| PPI Non-user | 1.38 (0.62-3.22) | 2.08 (0.85-5.25) |
| New PPI user | 1.14 (0.20-4.67) | 3.30 (1.11-9.73) |
| PPI Continued | Reference | Reference |
| Age | 1.00 (0.95-1.07) | 0.96 (0.93-0.99) |
| Post-discharge systemic antibiotics | 0.66 (0.06-3.62) | NA |
| Prior CDI | 26.0 (8.6-76.9) | NA |
| Active anticoagulant or antiplatelet use | NA | 1.51 (0.69-3.47) |
| Prior UGIB | NA | 6.82 (2.77-16.28) |
| Cirrhosis | NA | 3.41 (1.25-8.59) |

NA - Not applicable, variable not included in the model

### 30-Day UGIB

30-day post-discharge UGIB occurred in 30 (0.6%) of patients. Relative to continued PPI use, new use was associated with a 98.4% probability of increased odds of UGIB (aOR=3.30, 95%CrI=1.11-9.73); no probable association was identified for deprescribed (aOR=1.83, 95%CrI=0.43-6.38) or non-users (aOR=2.08, 95%CrI=0.85-5.25, **Table 2**).

## DISCUSSION

In this substudy of the MedSafer trial^10^, PPI desprescription was unexpectedly associated with an increased odds of CDI, but not UGIB. Furthermore, our study did not find a significant association between new PPI use and CDI.

Most evidence linking PPI use with CDI originates from observational studies and meta-analyses of such data^3^. Interpretation of these studies may be hindered by outcome and exposure misclassification, confounding by indication, protopathic bias, and selective reporting. CDI was not significantly higher in patients assigned to PPI versus placebo (P=0.18) in a trial of 18,000 patients with cardiovascular disease^13^.

Our analysis suggests deprescribing PPIs paradoxically increased the odds of CDI. This finding could reflect a transient disruption of the gut microbiome or bile acid metabolism following cessation of acid suppression, which may temporarily reduce colonization resistance and/or facilitate *C. difficile* proliferation^14^. Alternatively, the observed relationship could be due to the development of gastrointestinal symptoms or diarrhea occurring as a symptom of PPI withdrawal with the limitations of CDI polymerase chain reaction testing being unable to differentiate colonization versus infection^15^. Finally, the results could be solely due to residual confounding, resulting from a more targeted deprescription of PPIs in patients at highest risk for CDI. Further confirmatory studies are warranted. One potential approach is a targeted PPI deprescription trial in patients with CDI on unnecessary PPIs, which could be nested within a platform study.

This substudy utilizes data from MedSafer, the largest deprescription trial conducted to date^10^ and employs robust probabilistic statistical methods. That said, it is subject to certain limitations. First, because the MedSafer RCT’s randomization was at the cluster level, the possibility of imbalanced, unmeasured confounders cannot be excluded. Second, there is potential for outcome and exposure misclassification from the outcome being patient reported and, thus, unverifiable and PPI exposure status being ascertained at discharge rather than at outcome assessment, respectively. Third, our 30-day follow-up may miss later events, given most PPI studies on CDI follow to 90 days^2^. Fourth, despite the large sample size, the event rates for CDI and UGIB were low, which underpowers estimates. Finally, we considered any episode of CDI, whereas most evidence linking PPIs with CDI looks at recurrence^3^.

## CONCLUSION

Our study suggests that PPI deprescription may unexpectedly increase the risk of CDI in the 30-days following deprescribing. This challenges the existing paradigm. While unnecessary PPIs should still be avoided, the discontinuation of a PPI among hospitalized patients purely to reduce the risk of CDI is not supported by this evidence. These data are hypothesis-generating and further confirmatory and mechanistic studies are needed.

## Data Availability

All data produced in the present study are available upon reasonable request to the authors

